# Higher Vaccination Rate Predicts Reduction in SARS-CoV-2 Transmission across the United States

**DOI:** 10.1101/2021.11.14.21266325

**Authors:** Jacky Au

## Abstract

The severe acute respiratory syndrome coronavirus 2 (SARS-CoV-2) began proliferating widely throughout the world in late 2019/early 2020, creating a global pandemic and health crisis. Although vaccines became available to the public approximately one year after the onset of the pandemic, there still remains much hesitancy surrounding vaccination even two years into the pandemic. One key concern comes from reports of breakthrough infections among the vaccinated that show comparable levels of peak viral load as the unvaccinated, calling into question the ability of vaccines to slow or prevent transmission. Therefore young, healthy individuals who are at low risk of serious complications themselves have little incentive to receive a vaccine that they are not convinced will protect others around them. To address this important concern, this article analyzes COVID-19 incidence in the United States as a function of each state’s vaccination rate. Results show that states with higher percentages of fully vaccinated individuals report fewer new cases among the remaining unvaccinated population. These data add to accumulating evidence that COVID-19 vaccinations can indeed slow the spread of SARS-CoV-2, and are an important tool in society’s arsenal to put this pandemic behind us.

## Introduction

The severe acute respiratory syndrome coronavirus 2 (SARS-CoV-2) causes a respiratory disease known as coronavirus disease 2019 (COVID-19). Symptoms include fever, difficulty breathing, loss of smell or taste, and a host of other ailments ranging from minor to severe. This is an urgent global health crisis, resulting in over 251 million infections and over 5 million deaths worldwide, with the United States alone contributing roughly 1/6 of these numbers (46.7 million infections and 758,000 deaths). Fortunately, the recent development and distribution of vaccines has done much to curtail this issue, with initial clinical trials showing remarkable efficacy in preventing hospitalizations and deaths (1,2). In fact, a few months after mass distribution of vaccines began in the United States, many states began to relax lockdown regulations such as removing mask requirements in order to allow some semblance of pre-pandemic life to resume. However, with the recent surge of new mutations such as the Delta variant (3) and persistent vaccine hesitancy among a large portion of the population (4), this global health crisis is far from over. In fact, one prominent case in the highly vaccinated state of Massachusetts received national attention recently when an outbreak occurred in Barnstable county, despite the majority of infected individuals being fully vaccinated (5). Moreover, the Center for Disease Control (CDC) report on this case went on to find comparable levels of viral load in the nose and throat of vaccinated and unvaccinated individuals based on PCR cycle threshold values, which indicates the possibility that even vaccinated individuals carry a significant risk of transmitting the virus (5).

Based in large part on the results of this report, the CDC went on to adjust their guidance to re-recommend universal indoor masking. This series of news events, along with the CDC’s response, has caused understandable concern among many and has raised questions regarding the true extent to which vaccines prevent individuals from contracting and spreading the virus. Claims have even arisen among those opposed to or hesitant about vaccines that vaccination only serves to *increase* the spread of SARS-CoV-2 since vaccinated individuals are running around with comparable viral load to the unvaccinated but fewer symptoms to indicate the presence of infection. Despite the abundance of evidence that has since come out suggesting that the vaccinated actually have lower overall infection rates, both symptomatic and asymptomatic (6–8) as well as faster recovery times that shorten the window of infectiousness (9,10), such beliefs about vaccine ineffectiveness still persist and stymie mass vaccination efforts (11). Although evidence that vaccines protect the individual from severe illness and hospitalizations (1,2,8,12) is generally accepted and relatively non-controversial, there has been less focus on the effectiveness of vaccines in preventing transmission to others (but see 12). However, this is a critical and timely issue to address, especially for the young and healthy unvaccinated individuals who are at the lowest risk of serious health complications from SARS-CoV-2 but have the highest propensity to transmit to others (14).

The purpose of this report, therefore, is to conduct a large-scale, nation-wide analysis on the association between a state’s vaccination rate and the development of new COVID-19 cases (incidence) among the remaining unvaccinated population. To answer this question, I used empirical real-world data on COVID-19 incidence and vaccination rates provided by the CDC for each of the 50 states in the United States of America, as well as Washington D.C. For ease of readability, references to states or state-level analyses in the remainder of the manuscript will implicitly also include Washington D.C. Data were analyzed starting from June, approximately 1 month after vaccines became widely available to the general public in the United States, through the present (September 2021 at the time of writing). Furthermore, several important confounds were considered for inclusion in the model as controls (see Variable Selection in Methods), such as population density or willingness to comply with other pandemic policies that have been shown to reduce transmission such as staying at home (15,16) or wearing masks in public (17,18). Testing frequency was also considered, since states that conduct more random testing of asymptomatic individuals would be more likely to report higher overall case numbers. However, perhaps the most important variable to control was previous incidence during the same months in the preceding year. Since the same state-specific idiosyncrasies that contributed to case load last year, such as political attitudes (19), tourist hotspots, regional climate (20), demographics (21), and many others likely also contribute to case load this year, controlling for previous incidence allows for the model to account for the aggregated variation from these state-specific idiosyncrasies without the risk of overfitting the model by including each one individually.

The main hypothesis of these analyses is that states with higher vaccination rates will also report fewer new COVID-19 cases, which would lend support to the idea that getting vaccinated can protect others as well as oneself. However, there are two other possible outcomes as well. There could be no relationship, which may indicate that the effect of vaccination on community transmission is too small or non-existent to be detected through the noise of the myriad other nation-wide factors that contribute to viral transmission. Or there could even be a positive relationship such that more vaccinated states report *more* new cases rather than less. This would lend support to the idea often promulgated by vaccine skeptics that vaccination does nothing to slow down spread or infection, but does alter the behaviors of the vaccinated by masking symptoms and making them more likely to resume life as normal. To foreshadow the results, the main hypothesis ended up being supported, as elaborated further below.

## Methods

### Data Aggregation

#### Primary Variables

Data on vaccination rates (22) and COVID incidence (23), the primary independent and dependent variables of interest, respectively, were obtained using publicly available datasets from the CDC. Vaccination rate was defined as the percentage of a state’s population that was fully vaccinated at the beginning of a particular month. Incidence was defined as the proportion of new cases that arose in a particular state in a particular month per 100,000 unvaccinated individuals. Incidence for New York and New York City were reported separately in the CDC dataset, but were aggregated together in the current analyses to represent the incidence for the entire state of New York. A small, but unknown, number of the cases recorded in the CDC dataset consisted of breakthrough infections among the vaccinated. Since the CDC only tracks severe breakthrough infections that lead to hospitalizations or deaths, and do not report these at the state level, it is impossible to disaggregate the number of breakthrough infections from the total number of new cases each month within this dataset. I therefore estimated breakthrough rates based on partial data provided by the Kaiser Family Foundation (24). Unfortunately, these data are only limited to the first half of 2021, and are largely unavailable for the months of interest (June through September). Moreover, data from half the states are not reported. Thus, the number of breakthrough infections in the current analyses is only a crude estimate, extrapolating from the rate of breakthrough infections in the beginning of the year, and applying mean substitution to the states that did not report data. Although these assumptions are tenuous at best, I argue that this makes very little difference to the overall analyses, as the overwhelming majority of new cases reported to the CDC occurs among the unvaccinated. On average, only 1.7% of new reported cases occur among the vaccinated while 98.3% occur among the unvaccinated, based on the 25 states that reported data in the first half of 2021. Thus, I argue that any range of plausible breakthrough infection rates represents just a drop in the bucket when subtracted from the much larger pool of unvaccinated cases. To substantiate this argument, I provide sensitivity analyses where I run Monte Carlo simulations using randomly generated breakthrough infection rates for each state ranging from 0.4% to 12%, varying in increments of 0.01%. These boundaries were chosen based on doubling the minimum and maximum reported breakthrough infection rates, in order to account for recent increases in transmissibility due to the surge of the delta variant. These values also match with breakthrough infection rates reported in the literature, which range from 0.02% to 13.3% (25–34), but mostly skew towards the lower end especially when taken from a random sample rather than vulnerable groups such as health care workers. 10,000 simulations were run.

Finally, after calculating incidence in the preceding manner, a z-transformation was applied separately for each month. This transformation was critical in order to standardize values between months and make valid comparisons because of the surge in cases over time due to the increasing spread of the Delta variant. Without this transformation, a spurious correlation would occur in which incidence would appear to rise even as vaccination rates increase over the months.

#### Control Variables

Previous incidence from 2020 was calculated exactly as above, with the exception that cases were taken as a proportion of the entire population of a given state rather than the unvaccinated population since vaccines were not available then. The remaining control variables were extracted from various sources as follows. Population density, as well as the total population of each state, were extracted from World Population Review (35) using 2021 estimates, since the official US Census data for 2021 are not available at the time of writing. Testing frequency was measured as the number of PCR diagnostic laboratory tests performed in each state, expressed as a percentage of that state’s population. These data were obtained from the U.S Department of Health and Human Services (36). Mobility was determined via GPS data from mobile devices that tracked the number of individuals in each state who stayed home on a particular day, summed over the course of each month and expressed as a percentage of the total number of mobile phones tracked in that state. These data were extracted from the Bureau of Transportation Statistics (37). Finally, data on mask compliance were provided by the Delphi Research Group at Carnegie Mellon University, who in partnership with Facebook, administered massive daily surveys starting from September 2020 asking about whether an individual wore a mask most or all of the time in public over the last 5-7 days. On average, each day included roughly 3840 responses per state. The daily percentage estimates provided by the Delphi group were weighted by the demographic breakdown of a particular state in order to provide a representative sample from that particular state. I then weighted these daily estimates by the sample size of respondents on that day and averaged over the entire year from September 2020 to September 2021 in order to produce one time-invariant measure per state that reflected the general propensity to wear masks in public over the past year. This was done instead of calculating dynamic monthly estimates of mask use specifically for the months of June through September for two reasons. First, previous research has shown mask use to predict lower

COVID-19 incidence in 2020 (17,18), but mask use has dropped off considerably since the widespread availability of vaccines in 2021, especially among the vaccinated. Thus, this variable might be measuring different things in 2021 compared to 2020, and averaging over the entire year smooths out the signal. Second, doing so provided stronger correlations with the other variables, especially vaccination rate. A stronger correlation produces a more meaningful, and often more conservative, analysis because the regression partials out the correlated variable and estimates the unique variance of vaccination rate over and above mask usage.

### Variable Selection

Although all control variables were selected on strong theoretical grounds, care was taken to avoid overfitting the model with too many unnecessary variables. Thus, an inclusion criterion was set such that a variable must be significantly correlated with either the dependent variable (incidence) or the primary independent variable (vaccination rate) in order to be included in the final model (38,39). A test of variance inflation factors (VIF) was also carried out to rule out multicollinearity. All variables were averaged across months to test their correlations during this model selection process.

### Statistical Analysis

All statistical analyses were conducted using Stata version 13.0 (40). I first performed a series of simple ordinary least-squares regressions for each month from June, 2021 through September, 2021. The dependent variable was the untransformed COVID-19 incidence and the independent variable was vaccination rate. This analysis was conducted in order to establish the existence of a simple relationship between vaccination rate and incidence, uninfluenced by any other extraneous variables, as per previous recommendations (38).

After establishing this simple relationship across all months of interest, I re-analyzed the data with a single model, while controlling for potential confounds. To do so, I used a random effects panel regression using month-aggregated data as the within-unit estimator of time and state as the cross-sectional between-unit estimator, according to the following model:

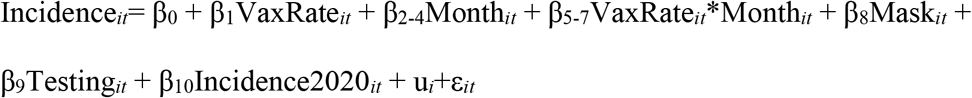

where the dependent variable, Incidence_*it*_, refers to the z-transformed incidence for state *i* during month *t*. β_0_ represents the overall regression intercept, β_1_ represents the coefficient for vaccination rate, β_2-4_ represent a 3×1 vector of coefficients for each month dummy variable, omitting the first one, June, β_5-7_ similarly represent a 3×1 vector of coefficients for the interaction of vaccination rate and each month dummy, β_8_ represents the coefficient for mask usage, β_9_ represents the coefficient for testing frequency, and β_10_ represents the coefficient for 2020 incidence. u_*i*_+ε_*it*_ represents the composite error term where u_*i*_ represents a random intercept for each state and ε_*it*_ is the idiosyncratic error of each state for each month. Hausman’s test (41) was used to test the existence of a correlation between u_*i*_ and the other independent variables in order to determine the appropriateness of the random effects model.

## Results

### Descriptive Statistics

Incidence and vaccination rates are reported in Table 1 for each state, along with overall summary statistics. Raw data for all other variables can be found in supplementary table S1.

**Table 1.** Incidence and Vaccination Rates per State

| State | June |  | July |  | August |  | September |  | Overall |  |
| --- | --- | --- | --- | --- | --- | --- | --- | --- | --- | --- |
|  | Vax % | Incidence | Vax % | Incidence | Vax % | Incidence | Vax % | Incidence | Vax % | Incidence |
| Alaska | 39.40 | 196.39 | 43.00 | 962.60 | 45.50 | 2937.09 | 47.30 | 6349.43 | 43.80 | 2611.38 |
| Alabama | 29.20 | 155.94 | 32.70 | 1206.35 | 34.40 | 4108.51 | 38.40 | 2325.37 | 33.68 | 1949.04 |
| Arkansas | 31.20 | 369.65 | 34.30 | 1792.63 | 36.50 | 3324.44 | 41.80 | 2357.70 | 35.95 | 1961.11 |
| Arizona | 36.10 | 262.89 | 43.10 | 711.64 | 45.30 | 1937.32 | 48.00 | 1965.04 | 43.13 | 1219.22 |
| California | 43.30 | 142.70 | 50.00 | 924.27 | 53.00 | 2142.41 | 56.00 | 1478.43 | 50.58 | 1171.95 |
| Colorado | 45.40 | 340.07 | 52.00 | 586.97 | 54.50 | 1490.16 | 57.00 | 1970.29 | 52.23 | 1096.87 |
| Connecticut | 53.70 | 112.00 | 60.80 | 386.74 | 63.40 | 1394.62 | 66.20 | 1435.52 | 61.03 | 832.22 |
| Washington D.C | 46.40 | 98.50 | 52.30 | 332.33 | 54.90 | 1442.73 | 57.40 | 1734.71 | 52.75 | 902.07 |
| Delaware | 43.40 | 153.31 | 50.10 | 333.15 | 52.80 | 1788.59 | 55.40 | 2938.70 | 50.43 | 1303.44 |
| Florida | 39.20 | 388.02 | 45.90 | 2420.71 | 49.00 | 5624.24 | 53.20 | 2943.48 | 46.83 | 2844.11 |
| Georgia | 32.10 | 143.46 | 36.70 | 698.36 | 38.70 | 3261.88 | 41.90 | 2735.90 | 37.35 | 1709.90 |
| Hawaii | 47.80 | 181.55 | 51.80 | 616.54 | 53.60 | 3119.46 | 55.40 | 2418.11 | 52.15 | 1583.92 |
| Iowa | 43.90 | 134.79 | 48.00 | 380.27 | 49.60 | 1621.21 | 51.80 | 3041.64 | 48.33 | 1294.48 |
| Idaho | 32.80 | 230.80 | 36.10 | 480.03 | 37.50 | 1752.02 | 39.30 | 3144.27 | 36.43 | 1401.78 |
| Illinois | 40.20 | 124.93 | 46.30 | 433.12 | 48.60 | 1543.70 | 51.30 | 1679.13 | 46.60 | 945.22 |
| Indiana | 35.50 | 220.32 | 40.00 | 443.32 | 44.30 | 2252.46 | 46.40 | 2826.21 | 41.55 | 1435.58 |
| Kansas | 38.50 | 207.87 | 42.00 | 894.31 | 45.30 | 2314.48 | 48.40 | 2549.65 | 43.55 | 1491.58 |
| Kentucky | 38.50 | 199.54 | 43.60 | 825.38 | 45.70 | 4166.96 | 48.70 | 4671.97 | 44.13 | 2465.96 |
| Louisiana | 31.30 | 296.43 | 35.00 | 2088.90 | 37.00 | 4866.52 | 41.60 | 1849.13 | 36.23 | 2275.25 |
| Massachusetts | 53.80 | 90.80 | 61.80 | 394.24 | 64.00 | 1567.03 | 66.00 | 2193.94 | 61.40 | 1061.50 |
| Maryland | 48.20 | 75.40 | 56.10 | 238.73 | 58.90 | 1165.37 | 61.60 | 1497.95 | 56.20 | 744.36 |
| Maine | 54.40 | 197.35 | 61.50 | 266.67 | 63.50 | 1163.08 | 65.90 | 3085.20 | 61.33 | 1178.08 |
| Michigan | 42.20 | 106.64 | 47.20 | 243.95 | 48.90 | 897.29 | 50.60 | 1833.62 | 47.23 | 770.37 |
| Minnesota | 46.20 | 125.35 | 52.00 | 270.69 | 53.90 | 1384.62 | 56.00 | 2393.18 | 52.03 | 1043.46 |
| Missouri | 34.40 | 493.27 | 39.20 | 1611.54 | 41.50 | 2152.96 | 45.20 | 1775.36 | 40.08 | 1508.28 |
| Mississippi | 27.10 | 190.41 | 29.80 | 1157.98 | 34.50 | 4696.25 | 38.40 | 2665.70 | 32.45 | 2177.58 |
| Montana | 38.30 | 278.49 | 42.90 | 421.71 | 44.40 | 1728.65 | 46.00 | 3933.12 | 42.90 | 1590.49 |
| North Carolina | 36.20 | 175.84 | 41.90 | 749.07 | 43.80 | 2842.75 | 46.50 | 2779.97 | 42.10 | 1636.91 |
| North Dakota | 36.60 | 152.22 | 38.90 | 196.39 | 40.20 | 1344.20 | 41.70 | 3252.19 | 39.35 | 1236.25 |
| Nebraska | 42.40 | 103.64 | 47.80 | 410.84 | 49.60 | 1637.14 | 52.10 | 2345.01 | 47.98 | 1124.16 |
| New Hampshire | 49.20 | 106.89 | 56.10 | 205.93 | 58.30 | 1189.26 | 59.70 | 2235.93 | 55.83 | 934.50 |
| New Jersey | 48.90 | 155.44 | 55.30 | 401.47 | 58.50 | 1425.71 | 61.60 | 1833.40 | 56.08 | 954.00 |
| New Mexico | 47.90 | 215.67 | 54.60 | 540.90 | 57.30 | 2400.65 | 60.10 | 2400.90 | 54.98 | 1389.53 |
| Nevada | 37.00 | 469.18 | 42.10 | 1254.00 | 44.50 | 1854.38 | 48.00 | 1822.73 | 42.90 | 1350.08 |
| New York | 47.00 | 121.20 | 54.20 | 407.58 | 57.20 | 1528.23 | 60.30 | 1902.35 | 54.68 | 989.84 |
| Ohio | 40.30 | 127.62 | 44.80 | 269.22 | 46.60 | 1439.69 | 48.50 | 3145.71 | 45.05 | 1245.56 |
| Oklahoma | 33.80 | 185.43 | 38.50 | 1058.53 | 40.30 | 2920.21 | 44.00 | 2695.51 | 39.15 | 1714.92 |
| Oregon | 45.50 | 300.54 | 53.80 | 584.08 | 56.00 | 2903.00 | 58.10 | 2934.74 | 53.35 | 1680.59 |
| Pennsylvania | 43.70 | 128.45 | 49.70 | 206.38 | 52.50 | 1120.21 | 55.20 | 2220.34 | 50.28 | 918.84 |
| Rhode Island | 51.80 | 131.80 | 59.00 | 477.70 | 61.60 | 2016.55 | 64.90 | 2564.42 | 59.33 | 1297.62 |
| South Carolina | 33.80 | 147.03 | 38.80 | 695.34 | 40.70 | 3664.97 | 43.50 | 3991.20 | 39.20 | 2124.63 |
| South Dakota | 42.40 | 65.67 | 45.50 | 168.78 | 47.00 | 1424.00 | 49.30 | 2759.02 | 46.05 | 1104.37 |
| Tennessee | 31.80 | 118.09 | 35.50 | 732.57 | 39.20 | 4243.67 | 42.00 | 3782.14 | 37.13 | 2219.12 |
| Texas | 35.60 | 229.04 | 41.30 | 758.80 | 43.90 | 2643.47 | 47.60 | 2793.09 | 42.10 | 1606.10 |
| Utah | 32.50 | 388.50 | 37.50 | 845.61 | 44.80 | 1652.95 | 47.80 | 2468.53 | 40.65 | 1338.89 |
| Virginia | 45.30 | 104.80 | 51.90 | 391.15 | 54.70 | 1859.22 | 57.40 | 2695.27 | 52.33 | 1262.61 |
| Vermont | 56.40 | 60.47 | 65.70 | 221.40 | 66.80 | 1498.93 | 67.90 | 2358.88 | 64.20 | 1034.92 |
| Washington | 46.70 | 351.84 | 54.70 | 653.45 | 57.70 | 2611.71 | 60.20 | 2987.04 | 54.83 | 1651.01 |
| Wisconsin | 44.60 | 97.07 | 49.60 | 342.28 | 51.80 | 1604.53 | 54.10 | 2789.16 | 50.03 | 1208.26 |
| West Virginia | 34.00 | 196.12 | 37.30 | 283.42 | 39.10 | 2049.96 | 39.70 | 4681.33 | 37.53 | 1802.71 |
| Wyoming | 31.80 | 492.11 | 34.50 | 761.62 | 36.70 | 2644.86 | 38.70 | 4267.96 | 35.43 | 2041.64 |
| Average | 40.94 | 198.85 | 46.34 | 661.56 | 48.78 | 2281.65 | 51.45 | 2696.07 | 46.88 | 1459.53 |
| (SD) | (7.26) | (110.43) | (8.60) | (485.15) | (8.52) | (1089.58) | (8.28) | (925.66) | (8.12) | (492.32) |

### Correlations between variables

Table 2 shows a pairwise correlation matrix between the dependent and all independent variables considered in the model. Population density and mobility were dropped from the final model due to a lack of significant correlations with both incidence as well as vaccination rates (*p’s* > 0.081). All other variables were retained.

**Table 2.**
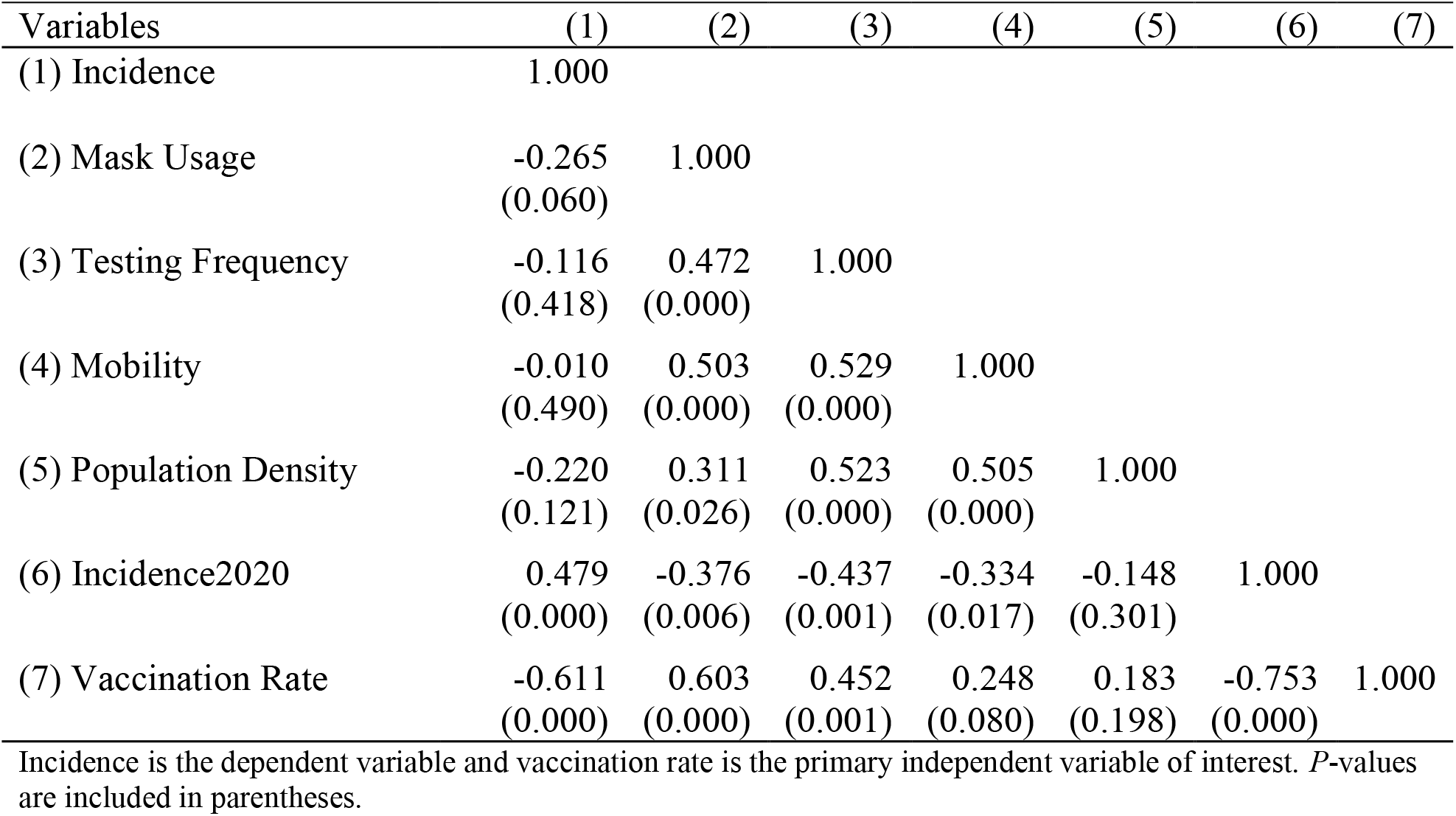
Correlation Matrix

| Variables | (1) | (2) | (3) | (4) | (5) | (6) | (7) |
| --- | --- | --- | --- | --- | --- | --- | --- |
| (1) Incidence | 1.000 |  |  |  |  |  |  |
| (2) Mask Usage | -0.265<br>(0.060) | 1.000 |  |  |  |  |  |
| (3) Testing Frequency | -0.116<br>(0.418) | 0.472<br>(0.000) | 1.000 |  |  |  |  |
| (4) Mobility | -0.010<br>(0.490) | 0.503<br>(0.000) | 0.529<br>(0.000) | 1.000 |  |  |  |
| (5) Population Density | -0.220<br>(0.121) | 0.311<br>(0.026) | 0.523<br>(0.000) | 0.505<br>(0.000) | 1.000 |  |  |
| (6) Incidence2020 | 0.479<br>(0.000) | -0.376<br>(0.006) | -0.437<br>(0.001) | -0.334<br>(0.017) | -0.148<br>(0.301) | 1.000 |  |
| (7) Vaccination Rate | -0.611<br>(0.000) | 0.603<br>(0.000) | 0.452<br>(0.001) | 0.248<br>(0.080) | 0.183<br>(0.198) | -0.753<br>(0.000) | 1.000 |

Incidence is the dependent variable and vaccination rate is the primary independent variable of interest. $P$ -values are included in parentheses.

Due to the significant correlations between the remaining variables, VIFs were tested in order to ensure multicollinearity was not present in the model. All variables had a VIF under 10 – Vaccination rate: 3.19, 2020 Incidence: 2.48, Mask Rate: 1.78, Testing Frequency: 1.43, Mean VIF: 2.22.

### Simple Linear Regression

Simple ordinary least-squares linear regressions were run separately for each month, regressing incidence on vaccination rate (Fig 1). In order to facilitate interpretation, the dependent variable was left untransformed in its natural units (cases per 100,000 unvaccinated people) for this analysis. All months showed significant negative associations: June (*b* = −6.530, *p* = 0.002, *r*^2^ = 0.184), July (*b* = −28.210, *p* < 0.001, *r* ^2^ = 0.250), August (*b* = −67.685, *p* < 0.001, *r* ^2^ = 0.280), and September (*b* = −44.010, *p* = 0.004, *r* ^2^ = 0.155).

**Fig 1.**
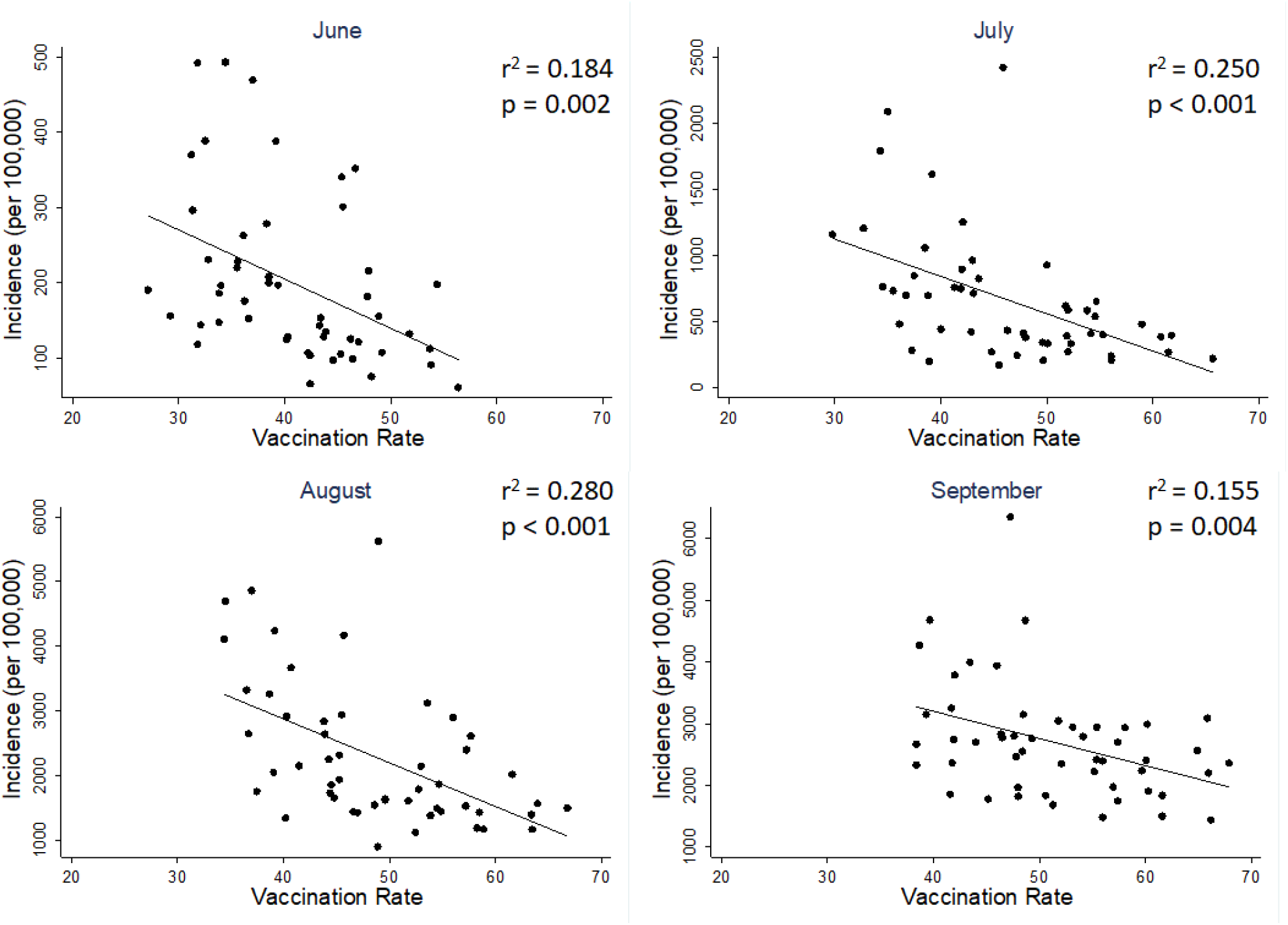
Simple Linear Regressions. Significant negative associations between vaccination rate and COVID-19 incidence were found during the months of June through September, with vaccination rate explaining between 15.5% to 28% of the variance in COVID-19 incidence. For ease of interpretation, the y-axis is unstandardized in these analyses, and left in its natural units (number of new cases per 100,000 unvaccinated people).

### Panel Data Regression

Panel data regression was used to confirm the effects of the simple linear regression. Hausman’s test detected no significant endogeneity (χ^2^(9)= 10.13, *p* = 0.340), and thus the random effects model was chosen over fixed effects (41). In concordance with the simple linear regression models, the random effects panel regression model showed a significant partial effect of vaccination rate (*b* = −0.055, *z* = −2.79, *p =* 0.005) referenced to the month of June, with no significant interactions for the months of July (*b* = 0.004, *z* = 0.18, *p* = 0.856), August (*b* = −0.0001, *z* = 0.00, *p* = 0.996), or September (*b* = 0.003, *z* = 0.14, *p* = 0.890), suggesting comparable effects of vaccination on COVID-19 incidence across all months. Previous incidence rates from 2020 were also predictive of current incidence rates (*b* =0.230, *z* = 2.73, *p* = 0.006), as was testing frequency (*b* = 0.028, *z* = 2.16, *p* = 0.031). The between R-squared value for the model was 0.399, suggesting almost 40% of the variation in COVID incidence between states can be explained by the variables in this model.

Given the lack of interactions with month, I re-ran the above regression without the interaction terms in order to arrive at a main effect of vaccination rate across all months: (*b* = −0.050, *z* = −3.51, *p <* 0.001). See Table 3 for all regression coefficients.

**Table 3.** Panel Regression

| *Independent Variables* | *b* | *SE b* | *p* |
| --- | --- | --- | --- |
| Vaccination Rate | -0.055 | 0.020 | 0.005 |
| July | 0.132 | 0.870 | 0.879 |
| August | 0.273 | 0.905 | 0.763 |
| September | 0.200 | 0.941 | 0.832 |
| Vaccination Rate X July | 0.004 | 0.020 | 0.856 |
| Vaccination Rate X August | 0.000 | 0.020 | 0.996 |
| Vaccination Rate X September | 0.003 | 0.020 | 0.890 |
| Incidence2020 | 0.230 | 0.084 | 0.006 |
| Mask Usage | 0.011 | 0.014 | 0.423 |
| Testing Frequency | 0.028 | 1.292 | 0.031 |

The dependent variable is COVID-19 incidence, z-transformed by month. Each independent variable is expressed as a percentage, so coefficients can be interpreted as standard deviation changes in response to a 1% change in the independent variable. The exceptions are Incidence2020, which is also z-transformed, and the month dummies which are coded as 1's and 0's. June was omitted as the reference.

### Sensitivity Analyses

Re-running the above panel regression using a range of different estimates for breakthrough infection rate did not change the results. Across 10,000 Monte Carlo simulations, the partial effect of vaccination rate for June ranged between −0.049 to −0.062 standard deviations, with all p-values showing statistical significance ranging from *p* = 0.005 to *p* = 0.033. None of the interactions with any other month were significant (all *p*’s < 0.71) with coefficients ranging from −0.001 to 0.007, suggesting comparable effects of vaccination rate across all months. The same held true looking at the main effect of vaccination rate by removing interaction terms. Over 10,000 simulations, the main effect of vaccination rate ranged between −0.044 to −0.056, with all p-values again in the significance range, from *p* < 0.001 to *p* = 0.002.

## Discussion

The present analyses provide compelling evidence for the real-world effectiveness of COVID-19 vaccines in reducing community transmission of SARS CoV-2 in the United States. Despite rising cases overall throughout the summer months due to the Delta surge and other factors, higher vaccination rates in a state at the beginning of each month still predicted fewer cases during that month relative to other states. Critically, COVID-19 incidence was calculated as a proportion of the unvaccinated population, not a proportion of the total population. Thus, these results go beyond the already plentiful evidence that the various COVID-19 vaccines are generally effective at protecting vaccinated individuals from symptomatic infection (1,2,8,12), but also provide evidence supporting the effectiveness of vaccines in protecting the surrounding community as well. Moreover, results were robust against a variety of different specifications for breakthrough infection rates, which were subtracted from the total number of new cases before calculating incidence among the unvaccinated. Although these breakthrough rates were not reported in the CDC dataset used in the current analyses and had to be roughly estimated, they occur so infrequently and are reported even less often, that their presence makes very little difference in the overall analyses. This was substantiated by Monte Carlo simulations that randomly generated breakthrough infection rates for each state using a range of realistic values and found that not one of the 10,000 simulations changed the interpretation of the results or had any substantial effect on the regression estimates (+/-0.006 standard deviations for the main effect of vaccination rate).

Throughout the observation period during the summer of 2021 (June through September), each percentage increase in a state’s vaccinated population was associated with a reduction in new COVID-19 cases by approximately 0.053 standard deviations. To illustrate the real world implications of this, Table 1 shows that one standard deviation throughout these months averaged to about 492.32 infections per 100,000 unvaccinated people. Thus, in a hypothetical population of 100,000 unvaccinated people, each 1,000 (or 1%) of them who became fully vaccinated at the beginning of June would be associated with an average of 26.09 fewer cases *per month* among their unvaccinated peers. By the end of September, these same 1,000 vaccinations would be associated with 104.37 fewer infections overall. More impressively, these results span a window of time during which the Delta variant was the predominant strain in the United States, suggesting that vaccines may be effective at slowing spread even against this highly infectious strain.

Of course, the problem inherent with observational data is the lack of randomization to a treatment or experimentally-designed placebo group, without which it is difficult to make causal claims about the association between increased vaccination rate and decreased COVID-19 incidence. However, these findings did not change even in the presence of a variety of important controls, including mask usage, testing frequency, and previous incidence. Therefore, alternative explanations relating to these variables can be ruled out. Most importantly, including previous incidence in the model serves as a powerful control because it effectively accounts for the aggregated variation due to state-specific idiosyncrasies that may influence the endemic spread of SARS-CoV-2 within a particular state. For example, a state that reported a high case load last summer due to large gatherings at popular beaches may also report a high case load this year due to the same beaches. Controlling for previous incidence accounts for the aggregated influences of these state-specific factors. In fact, Table 2 shows a fairly strong correlation between previous and current incidence (r=0.48, *p* < 0.001), supporting the existence of these state-specific factors. This is a particularly important consideration given that a random-effects model was used, which favors more precise coefficient estimates compared to a fixed effects model, but at the expense of possibly introducing some level of omitted variable bias due to correlations between the unit-level (i.e., state) error terms and the other independent variables (42). Controlling for state-level idiosyncrasies partially mitigates this issue by reducing variation associated with state-level error, achieving some of the same goals as a fixed effects model in terms of offering some level of correction against omitted variable bias while maintaining the improved precision of a random effects model.

An additional and important consideration is that summer incidence during 2020 was related not only to summer incidence during 2021, but also had a strong negative correlation with the primary independent variable, vaccination rate (*r* = −0.75, *p* < 0.001). Therefore, increased vaccination rate predicted lower incidence both this year and last. Ostensibly, this is problematic since vaccines clearly cannot have a retroactive effect; therefore, this independent variable must reflect more than just vaccination rate, but perhaps also the willingness to employ a constellation of other pandemic policies as well that may have influenced transmission over the last year. For example, Table 2 shows that states with higher vaccination rates this summer also wore masks more often throughout the last year (*r* = 0.603, *p* < 0.001), indicating that a part of the variation in vaccination rate (*r*^2^ = 0.364) overlaps with the variation in mask usage. Although there are likely other unmeasured variables at play, this is one important factor that could mediate the retrospective relationship between vaccination rate and 2020 incidence, especially given that mask usage has already been shown to predict lower COVID-19 incidence in 2020 (17,18). There are two important takeaways from all this. First is that vaccines are not the only tool in society’s arsenal to stem the tide of the pandemic. Although the present results say little about which specific policies and practices, other than vaccination, are helpful, they do suggest that states that tend to favor vaccination also tend to favor other recommendations and guidelines, and ultimately tend to fare better. Second, it is important to bear in mind that these correlations do not take away from the main message of the manuscript, which is that vaccinations, specifically, predict lower COVID-19 incidence. Rather, the results of the panel regression show that the effect of vaccination rates is robust enough to still uniquely predict current incidence, even above and beyond its relationships with mask usage and previous incidence.

Furthermore, these findings are in line with recent studies that show fewer symptomatic *and* asymptomatic infections among the vaccinated, as assessed by routine laboratory PCR testing (6–8,43). Thus, it’s no surprise that states with a more vaccinated populace seem to offer more protection from infection, even to its unvaccinated residents. In addition, evidence also shows that aside from being less likely to host the virus in the first place, vaccinated individuals with breakthrough infections also carry less overall viral load throughout the duration of infection, despite having similar peak levels as the unvaccinated at the beginning of infection (9,10,44). Moreover, even for the same levels of viral load, less infectious virus was found in respiratory samples among the vaccinated, indicative of less viral shedding (45,46). Therefore, several mechanisms seem to be at play that limit the spread of virus from the vaccinated. Aside from the current study, this has also been borne out empirically in other recent large-scale studies in Israel that showed reduced transmission from vaccinated individuals to their households compared to the households of unvaccinated individuals (13,47).

In totality, the evidence that COVID-19 vaccinations reduce the spread of SARS-CoV-2 is quite strong and consistent. Although the public messaging from the government and other authorities so far has focused primarily on the effectiveness of COVID-19 vaccines in protecting the individual, enough evidence has accrued now that this message should shift to the effectiveness of vaccines in protecting the community. This message would be especially pertinent to young, healthy adults who are the least likely to suffer major complications if infected, and therefore have the least personal incentive to get vaccinated. Although the vaccination of older adults has done much to reduce hospital burden and save lives among the elderly, it is now the young and healthy that need to be prioritized for vaccination. In support of this, an enterprising study by Monod et al. (14) modeled infection dynamics across the nation and correlated this to cell phone mobility data. They found that the contact patterns of younger adults, aged 20-49 were most predictive of COVID-19 transmission and deaths than any other age group. Although vaccine mandates may be one viable solution to increase vaccination rates among younger adults across the country, this has been met by severe pushback from many, especially by those who consider vaccination to be solely a personal health choice. Considering that the strongest and most publicized evidence of vaccine effectiveness so far has been the reduction of hospitalizations and deaths among the individual (1,2,6,8,12,48), and that the public health narrative from the government and CDC has accordingly focused on this, it is perhaps not surprising that many would cling to this personal choice argument. However, based on the present results as well as accumulating evidence from the literature of reductions in infectiousness and reductions in community transmission, it may be time to refocus this narrative to recognize that the benefits of vaccination are not *solely* personal, but also communal as well.

## Data Availability

All data produced in the present work are available in the supplementary materials.

## Acknowledgments

I would like to thank Dr. Nancy Tsai (Massachusetts Institute of Technology), Dr. Carley Karsten, Dr. Laura Berkowitz (Cornell University), and Dr. Susanne Jaeggi (University of California, Irvine) for their valuable scientific input and discussions during the early stages of the manuscript, as well as for the encouragement to publish. I am supported by the National Institutes of Health, through Grant TL1TR001415. I declare no financial or other conflicts of interest.

